# Protocol for: A Simple, Accessible, Literature-based Drug Repurposing Pipeline

**DOI:** 10.1101/2024.07.18.24310641

**Authors:** Maximin Lange, Meredith Martyn, Eoin Gogarty, Philip Braude, Feras Fayez, Ben Carter

**Affiliations:** Artificial Intelligence in Mental Health Lab, Department of Psychosis Studies, Institute of Psychiatry, Psychology and Neuroscience, King’s College London; Biostatistics & Health Informatics Department, Institute of Psychiatry, Psychology and Neuroscience, King’s College London; Department of Psychosis Studies, Institute of Psychiatry, Psychology and Neuroscience, King’s College London, South London and Maudsley NHS Foundation Trust, King’s College Hospital NHS Foundation Trust; Consultant Geriatrician, North Bristol NHS Trust; Imperial College Healthcare NHS Trust; King’s Clinical Trials Unit, Institute of Psychiatry, Psychology and Neuroscience, King’s College London

## Abstract

We will develop a novel approach to drug repurposing, utilising Natural Language Processing (NLP) and Literature Based Discovery (LBD) techniques. This will present a simplified, accessible drug repurposing pipeline using Word2Vec embeddings trained on PubMed abstracts to identify potential new medications to be repurposed. We present this approach in the context of antipsychotics, but it could be repeated for any available medication.

The research is structured in three stages:

1. Identification of candidate medications using Word2Vec algorithm trained on scientific literature.
2. Empirical testing of identified candidates using a large hospital dataset to explore protective effects against disease onset.
3. Validation of findings using a second, independent dataset to assess generalizability.

This method addresses limitations in current machine learning-based drug repurposing approaches, including lack of external validation and limited accessibility. By leveraging Word2Vec’s ability to capture semantic relationships between words, the study aims to uncover hidden connections in medical literature that may lead to novel therapeutic discoveries.

The protocol emphasizes transparency and reproducibility, utilizing publicly available electronic health record (EHR) databases for validation. This approach allows for tangible results even for researchers with limited machine learning expertise, bridging the gap between biomedical and information systems communities.

## Key Study Contacts

**Chief Investigator:** Maximin Lange

**Study Senior Methodologist:** Prof Ben Carter

Sponsor Joint-sponsor(s)/co-sponsor(s): **N/A**

**Funder(s):** No specific funding was obtained for the delivery of this study. Maximin Lange is supported through a studentship by the London Interdisciplinary Social Science Doctoral Training Partnership (LISS DTP). Ben Carter is supported through the NIHR Maudsley Biomedical Research Centre at the South London and Maudsley NHS Foundation Trust in partnership with King’s College London

**Key Protocol Contributors:** Maximin Lange, Ben Carter, Meredith Martyn, Phil Braude

## Study Summary

Study Title:

A Simple, Accessible, Literature-based Drug Repurposing Pipeline –

Validating*.J*predictions using MIMIC-IV, BRATECA and CRIS-SLaM platforms

Internal ref. no. (or short title): Simple, Literature-based Drug Repurposing

Study Design: Retrospective observational database study

**Study Participants:** Patient data held within the Medical Information Mart for Intensive Care IV (MIMIC-IV), Brazilian Tertiary Care (BRATECA) and Clinical Record Interactive Search (CRIS) database from Hospitals in Boston, MA, USA, 10 hospitals in two Brazilian states and the South London and Maudlsey NHS Foundation Trust (SLaM) in London, UK.

Follow up duration (if applicable): No follow-up

**Planned Study Period:** MIMIC IV: 2008-2019, BRATECA: 2020-2021, CRIS: 2006-2024.

Research Question/Aim(s):

- **To develop a simplified drug repurposing pipeline utilizing Word2Vec embeddings in the context of literature-based discovery, applied to the example of psychosis, but usable in any medical condition.**

This aim focuses on the core methodology of step 1 of the project, emphasizing a streamlined, easy to navigate use ofWord2Vec embeddings and cosine similarity in this specific context.

**•** To demonstrate the validity of the novel drug repurposing pipeline by evaluating protective effects of newly proposed medications against psychosis onset in real-world patient populations.

This aim emphasizes the practical application of the research, which is step 2 of the project, moving from theoretical methodology to tangible results. It specifically mentions the use of real-world data (hospital datasets) to validate the findings, which adds a layer of clinical relevance to the research.

**•** To assess the generalizability and robustness of the Word2Vec-based drug repurposing pipeline by replicating the evaluation of identified drug candidates in multiple independent datasets.

This aim addresses the important issue of reproducibility and external validation of research findings; step 3 of the project. By replicating the analysis in different datasets, it seeks to confirm the reliability and potential clinical significance of the identified drug candidates.

## Plain Language Summary

We will develop a method to identify drugs that might be beneficial for treating other diseases than those for which they were developed. We will present an application of this in psychosis and related disorders, which are mental health conditions with symptoms creating a disconnect from reality; sufferers often see or hear things that are not there.

We will use a machine learning technique, called Word2Vec, that helps machines understand word meanings. This technique converts words into numbers that represent their meaning. In our model, each word becomes a point in a 300-dimensional space, with similar words positioned closer together.

Word2Vec has been used to make new discoveries from already published literature. In materials science, it was trained on scientific papers, and it learned about chemical elements and their properties. It suggested new materials with specific properties that hadn’t been previously considered.

By analysing medical literature, Word2Vec might uncover drug-disease connections that human researchers have not identified. In this study, we will use Word2Vec to search for potential new psychosis treatments. We will train our algorithm on medical papers and ask it to suggest drugs that might be effective, even if they have not been directly linked to psychosis before.

We anticipate this approach could speed up treatment discovery by highlighting unexplored directions. It would demonstrates how simple artificial intelligence tools can assist in solving complex problems in medicine, even without extended training in these methods.

In the question generating first stage, our method will propose drugs to be repurposed, which are already approved drugs for other medical conditions and unrelated to psychosis. We will search within the three databases, out of the top 20 medications suggested by our method, the one to have the largest number of patients with a diagnosis of psychosis or related disorders across the datasets. This medication will be chosen for further examination.

In the second stage we will test the hypothesis that our medication could offer a protective effect against progression to psychosis or related disorders using an independent data source. Our study will look at adults admitted to hospital at least once. The first time, for something unrelated to psychosis or related disorders, at which point they received our medication. Following discharge, some were readmitted for psychosis or related disorders, while some were not. We will compare those that received our medication to those that didn’t to see if there was any association with the risk of diagnosis (or treatment) for psychosis or related disorders. To test this hypothesis, we will use a large US hospital dataset (MIMIC-IV; Johnson et al., 2023).

Then, within stage 3 we will repeat stage 2 using a second independent data source we will validate the findings from stage 2 to see if a protective effect may exist elsewhere, in datasets from Brazil (BRATECA; Dias et al., 2022) and the UK (CRIS; Stewart et al., 2009).

Our results will help researchers find new drugs for psychosis and related disorders faster. Ultimately, these will aid clinicians and health systems to improve the care for people with these illnesses. If our methodology proves feasible and helpful, this could be repeated for other medications and illnesses.

## Funding and Support in Kind

FUNDER(S)

London Interdisciplinary Social Science Doctoral Training Partnership

NIHR Maudsley Biomedical Research Centre at the South London and Maudsley NHS Foundation Trust in partnership with King’s College London

Role of Study Sponsor and Funder

Financial. Not directly for this study.

Study Group

Maximin Lange

PhD Student in Artificial Intelligence in Mental Health, King’s College London Dr Meredith Martyn

Medical Statistician. Department of Biostatistics and Health Informatics, Institute of psychiatry, psychology and neuroscience, King’s College London.

Eoin Gogarty

Data Scientist, Department of Psychosis Studies, Institute of Psychiatry, Psychology and Neuroscience, King’s College London, South London and Maudsley NHS Foundation Trust, King’s College Hospital NHS Foundation Trust

Prof Ben Carter

Professor of Medical Statistics, Department of Biostatistics and Health Informatics, Institute of psychiatry, psychology and neuroscience, King’s College London.

Dr Philip Braude

Consultant geriatrician, North Bristol Trust, Bristol, UK Dr Feras Fayez

Medical Doctor, Imperial College Healthcare NHS Trust, London, UK

Role Responsibility

All members have responsibility for study development and delivery

Protocol Contributors

Prof Ben Carter

Maximin Lange

Dr Meredith Martyn Dr Philip Braude Eoin Gogarty

Dr Feras Fayez

**Role**

Professor of Medical Statistics, King’s College London

PhD Student in Artificial Intelligence in Mental Health, King’s College London Medical Statistician, King’s College London

Data Scientist, King’s College London, South London and Maudsley NHS Foundation Trust, King’s College Hospital NHS Foundation Trust

Consultant geriatrician, North Bristol Trust, Bristol, UK

Medical Doctor, Imperial College Healthcare NHS Trust, London, UK

Responsibility

Wrote the protocol. Writing of manuscript and dissemination Wrote the protocol. Writing of manuscript and dissemination

Wrote the protocol. Analysis of data. Writing of manuscript and dissemination Curation of data, contributed to the manuscript

Contributed to the protocol

Contributed to the protocol, contributed to the manuscript

## Research in context

### Evidence before this study

We searched, PubMed, Scopus, MEDLINE, IEEE and Web of Science databases for all studies published from database inception to Jul 15, 2024, using the following search terms: [(“word embed*” OR “Word2Vec” OR ““NLP” OR “natural language proces*) AND ((“drug” OR “medication” OR “pharmacol*”) AND (“discovery” OR “repurpos*” OR “reposition*” OR “develop*”))]. We found over thirty reviews discussing concepts related to literature-based discoveries (LBD) or general machine learning (ML)-driven drug repurposing systems. All relevant references were checked for additional citations. The reviews establish LBD as a reliable method of biomedical knowledge creation and hypothesis generation. Drug repurposing candidates were at times being validated by external Electronic Health Records (EHR) datasets; no paper reported validation on more than one external EHR dataset. We note that drug repurposing pipelines often rely on complicated techniques to identify candidates. Simpler methods, closely related to ours, have been presented but were never validated using external datasets.

As such, drug repurposing pipelines utilising machine learning methods remain heavy-handed and not sufficiently validated.

### Added value of this study

We report a comprehensive, accessible method to identify medication candidates for repurposing.

This method will be validated using three external, heterogenous datasets from three different health systems; all of them are publicly available. This method can be made accessible to researcher without extended machine learning knowledge or training.

### Implications of all the available evidence

Our findings investigate whether the Word2Vec algorithm trained on large quantities of PubMed abstracts can be used to identify drugs with potential antipsychotic properties. This method could then be repeated for other available drugs and medical conditions. This method would have the potential to point researchers in the most promising directions; directions which would otherwise might never have occurred to the community.

Future studies should improve model training and utilise more external EHR databases for external validation. A web application that opens the technique up to non-ML-experts is needed.

## 1 Background

### Literature based discovery

Machine learning techniques have gained prominence in drug repositioning and repurposing (For reviews see Chen et al., 2018; Gupta et al., 2021; Zong et al., 2022; Koshechkin et al., 2022; Xue et al., 2018; Yang et al., 2022; Dara et al., 2022; Carracedo-Reboredo et al., 2021; Stephenson et al., 2019; Wang et al., 2024; Ghandikota & Jegga, 2024; Pan et al., 2022).

A sub-discipline of this field is the utilisation of Natural language processing (NLP), an area within artificial intelligence that enables information systems to understand, interpret, and generate human language in a way that is both meaningful and contextually relevant. NLP methods use deep neural networks to extract implicit, dormant patterns present in large scientific text corpora, galvanising Literature Based Discovery (LBD; Lardos et al., 2022; Gopalakrishnan et al., 2019; Trajanovet al., 2023; Bhatnagar et al., 2022; Crichton et al., 2020).

LBD has been conducted for almost forty years (Smalheiser et al., 2023; Henry & McInnes, 2017; Hristovski et al., 2013; Ganiz et al., 2005), initially not widely adopted outside of computer science (Lardos et al., 2022; Henry & McInnes, 2017), while gaining prominence as of recent (Cesario et al., 2024). Translation into mainstream drug research has primarily been hindered by limited empirical evaluation of new discoveries, lack of communication between biomedical and information systems communities and computationally intense nature of involved methods (Moreau, 2023; Cheerkoot- Jalim & Khedo, 2021; Henry & McInnes, 2017; Lardos et al., 2022; Gupta et al., 2021).

These shortcomings are increasingly being addressed by developers (Cesario et al., 2024; Zong et al., 2022; Henry & McInnes, 2017). Still, LBD drug repurposing pipelines are seldom validated with external EHRs, and if done, only on one single dataset and not sufficiently transparent (Zong et al., 2022; Lardos et al., 2022; Xue et al., 2018; Dara et al., 2022; Ji et al., 2020; Cheerkoot-Jalim & Khedo, 2021). In cases where EHRs are utilised, circulation to wider audiences is hindered through limited accessibility and understanding of EHRs (Zong et al., 2022).

### Word2Vec Algorithm

Word2Vec (Mikolov et al., 2013) is a Natural Language Processing (NLP) technique that mathematically models relationship between words. This happens by converting them into vectors, i.e., numerical representations of words that capture information about the word meaning (Johnson et al., 2024). Word2Vec takes each of the distinct words from a predefined text corpus and converts them into a 300-dimensional vector, in other words an array of 300 numbers.

The algorithm uses the numbers to identify how they are related to other numbers, i.e., in which way words are related to each other. These vectors represent words in a multi-dimensional space where words with similar meanings are placed closer together. By analysing large amounts of text data, Word2Vec learns to predict the context of a word based on its neighbouring words. In simple terms, Word2Vec helps computers learn the meaning of words by evaluating how they are used in a text.

Using standard mathematics, Word2Vec can subtract vectors. An algorithm trained on regular journalistic text, can take the vector resulting from ‘king’ subtracting it from that of ‘queen’, and get the same result as when subtracting the vector for ‘man’ from that of ‘woman.’ This is possible without human supervision (Di Gennaro et al., 2021).

Instead of journalistic text, Tshitoyan et al., (2019) trained the Word2Vec algorithm on abstracts from materials science literature. The algorithm grasped the meaning of scientific terms and concepts e.g., crystal structures of metals, only by realising positions of words within the abstracts in relation to other words. In the same way as the lay-text-trained Word2Vec algorithm can correctly solve “king – queen + man = woman”, Tshitoyan’ et al.’s (2019) Word2Vec algorithm was able to solve “ferromagnetic – NiFe + IrMn” correctly with “antiferromagnetic”. When vectors for chemical elements were projected onto two dimensions, the algorithm could further grasp relations between elements on the periodic table.

### Word Similarities Uncover Hidden Knowledge

The most important contribution made by Tshitoyan et al. (2019), however, was their finding of a substantial number of materials with a high cosine similarity to the word ‘thermoelectric’, which never appeared explicitly in the same abstract with this word, or any other words that would identify a material as thermoelectric. Instead of treating this as noise or hallucinations of the model, these materials were further examined for potential novel thermoelectric properties. The ten most promising candidate were found in experimental testing to indeed exhibit unusually high thermoelectric properties. This demonstrated that the Word2Vec algorithm, when trained on scientific texts, can be leveraged to address gaps in materials science research which are hard to access for human scientists.

### Hidden Medical Knowledge

We believe this approach can be leveraged to make yet unbeknownst connections which aid therapeutic discoveries in human medicine.

## 2 Rationale

### Overview

To overcome the mentioned limitations of other ML-based drug repurposing methods, we present a simple and accessible LBD drug repurposing method, tangible even for researchers with limited knowledge in machine learning. Our validation EHR databases are publicly available.

While our method is applicable to more than one disorder/medication, we focus here on finding new psychiatric medications.

We will use a machine learning algorithm to create the list and generate the hypothesis of repurposed medication. We will then test this theory and validate using two separate external datasets to assess the evidence of repurposed medications offering a protective effect.

## 3 Research Question/Aims

**•** To develop a simplified drug repurposing pipeline utilizing Word2Vec embeddings in the context of literature-based discovery, applied to psychosis and related disorders.

This aim focuses on the core methodology of step 1 of the project, emphasizing a streamlined, easy to navigate use ofWord2Vec embeddings and cosine similarity in this specific context.

**•** To test the novel drug repurposing algorithm by evaluating protective effects of newly proposed medications to delay onset of specific illness, in real-world patient populations.

This aim emphasizes the practical application of the research, moving from theoretical methodology to tangible results. It specifically mentions the use of real-world data (hospital datasets) to validate the findings, which adds a layer of clinical relevance to the research.

**•** To assess the generalizability and robustness of the Word2Vec-based drug repurposing pipeline by replicating the evaluation of identified drug candidates (and drug class) in multiple independent datasets.

This aim addresses the important issue of reproducibility and external validation of research findings. By replicating the analysis in different datasets, it seeks to confirm the reliability and potential clinical significance of the identified drug candidates.

## 4. Objectives

### Objective

Whether the Word2Vec algorithm trained on large quantities of PubMed abstracts can be used to identify drugs with potential antipsychotic properties. This method could then be repeated for other available drugs and medical conditions. This method would have the potential to point researchers in the most promising directions; directions which would otherwise might never have occurred to the community.

### Stages

Stage 1: Identify candidate medications

Identifying medications to be repurposed for psychosis and related disorders

Stage 2: Test the candidates using empirical data.

Test the theory by exploring if the drug (and expanded to class) of medications offer any protection against disease onset using a large hospital dataset in a population without the condition at onset.

Stage 3: Validate the findings of stage 2 in a second, independent data source.

Repeat Stage 2, to assess if the pattern occurs on a second independent dataset

## 5 Study Design, Methods of Data Collection and Data Analysis

Our methodology can be split into three parts:

1. Hypothesis Generation / Training the algorithm and identifying repurposed drugs relevant to psychosis
2. Find empirical evidence for efficacy of newly suggested medication
3. Validate part 2) on an independent data source

### Stage 1 - Hypothesis generation

#### Machine Learning Methods

##### Training the algorithm

###### Corpus creation

The National Library of Medicine has made the whole of PubMed downloadable free of charge at https://ftp.ncbi.nlm.nih.gov/pubmed/baseline/.

In its entirety, the file would be large (∼360GB), making it unrealistic to handle and hard to use in its entirety given our computing resources. Hence, we will train our model on a as large a chunk of it as is possible. Details on our data curation will be disclosed in the manuscript.

The abstracts will be uploaded into Google Colab and cleaned. Non-alphabetic characters and extra spaces removed, capitalized words with more than four characters removed (most abstracts contained words like BACKGROUND or METHODS etc.). All numbers and punctuation will be removed as well as single letter words and the word ‘copyright’. Finally, stop words will be removed. Stop words are a set of commonly used words in a language that are often excluded from text processing and analysis, such as “a,“ “the,“ “is,“ “are,“ etc. These words carry little useful information and are typically removed during natural language processing and text mining to focus on more meaningful content (Sarica & Luo, 2021). The whole corpus will be made available.

###### Word2Vec Training

We will use a modified Word2Vec implementation in gensim (https://radimrehurek.com/gensim/) running on Google Colab to generate the word vector representations.

Hyperparameters will be found through optimization for performance on 60 ground truths (30 medical, 30 grammatical). The code used for the training and the full list of analogies used for optimisation will be made available.

##### Identifying drugs to be repurposed

We will screen two medications to get new candidates for treatment options, amisulpride and clozapine. We chose amisulpride as it is the alphabetically first commonly prescribed antipsychotic (Royal College of Psychiatrist, 2022) and clozapine as it is the only available treatment for treatment resistant schizophrenia (Maudsley Prescribing Guidelines, 2021).

A new prediction will be every word close to the input word, which is a medication and has not been mentioned together with the following terms in a PubMed abstract: (schizo* OR psychot* OR psychosis OR antipsychotics OR amisulpride/clozapine) between 2000-2023. We chose this time range to allow for fewer abstracts to be chosen, not all of PubMed, so potential limits in computational resources can be overcome.

In cases where there will be five or less abstracts in which a word and the above query were mentioned together, we will check if the abstracts in question are featured in our training corpus. This will always be a possibility, since, as described earlier, we likely only put a fraction of all PubMed abstracts into the model. If the abstracts will not have featured in the training corpus, the word will be considered a prediction, since the model would not have known about a connection of the two words through abstracts.

Choosing which newly suggested medication to investigate for protective properties We will search in MIMIC-IV, as it is our most accessible dataset, which of the top 20 newly suggested medications has the most likely of a positive diagnosis for the condition under investigation at some point in the future.

### Stage 2 - Hypothesis testing algorithm on external dataset

#### Statistical methods

The analysis will involve three data sources. A testing dataset will use the MIMIC-IV database. The validation datasets will use the BRATECA and CRIS data sources.

Retrospective observational cohort study using the MIMIC-IV database from 2008 to 2019, BRATECA database from 2020-2021 and CRIS database from 2006-2024. Data will be anonymously transferred to King’s College London devices for analysis. Statistical methods have been developed by two statisticians (BC and MM).

The medication class discovered by the ML algorithm will be tested using an external data. The participants will be determined as having the medication, compared to those not receiving the medication at the index hospitalisation.

Eligibility Criteria Medication Group Inclusion Criteria:

1. Index Hospital Admission: The patient’s initial documented admission to the hospital
2. Medication(s) Administration: The patient received our target medication(s)
3. Age: 18 years or older at the time of their first admission.
4. No Prior related Disorder using ICD 9 / 10 codes: 290-299 / F01-09, 20-29

We are very broad with what counts as psychosis or related disorders, i.e. we are including endogenous, non-organic psychoses as well as exogenous, organic psychoses. This is since our algorithm will suggest medications close to antipsychotics, and antipsychotics are being prescribed for all these conditions (Prescribing Observatory for Mental Health, 2017).

Exclusion Criteria:

1. Prior Related Disorder using ICD codes
2. Medication(s) not during index Admission: The patient did not receive medication(s) target during their initial hospital stay.
3. Age: The patient was under 18 years old at the time of their first admission.
4. Prior related Disorder

Control Group

Inclusion Criteria:

1. Index Hospital Admission: The patient’s initial documented admission to the hospital.
2. Medication(s) Administration: The patient did not receive our target medication(s) during their first admission.
3. Age: 18 years or older at the time of their first admission.
4. No Prior Related Disorder Exclusion Criteria:
5. Prior Related Disorder
6. Medication(s) during index Admission: The patient did receive target medication(s) during their initial hospital stay.
7. Age: The patient was under 18 years old at the time of their first admission.
8. Prior related Disorder

#### Statistical Analysis

Analysis will be conducted using the MIMIC-IV Clinical Dataset with a time-to-event endpoints. The index timepoint will be a patient’s first hospitalisation, with the event of interest as time to second- hospitalisation due to psychosis or related disorders. The two time-to-event curves between patients who have been prescribed our target medication, identified in stage 1 of this protocol.

We will extend the test of our theory by exploring if the class of medications are protective against disease onset using a large hospital dataset in a population without the condition at onset.

Primary exposure: Class of medications that the medication suggested by our algorithm belongs to.

Secondary exposure: Specific medication suggested by our algorithm

Primary outcome: Time from discharge to hospital admission with for illness or Disorder.

#### Primary outcome analysis

A Cox proportional hazard regression model will be used to estimate the hazard ratio (HR). If the two-sided p-value is greater than this significance threshold (α) of 5%. We will adjust for patient age and sex alongside the class of drug. We will estimate the adjusted HR (aHR) and present it alongside the p-value. If there is not adequate evidence against the null hypothesis. This would suggest that patients prescribed our target medication(s) do not have a different hazard rate to second- hospitalization due to psychosis or related disorders than patients who are not.

### Stage 3 – Validation on a second external dataset

Analysis performed for hypothesis testing using the MIMIC-IV Clinical Dataset will be repeated using the datasets BRATECA (Brazilian Tertiary Care Dataset) and CRIS (Clinical Record Interactive Search). BRATECA provides clinical data on 10 Brazilian hospitals, and CRIS provides clinical data from the South London and Maudsley NHS Foundation Trust in the UK.

As the clinical data available is not consistent across the datasets, the analysis will be different to the analysis conducted in hypothesis testing. Instead, the outcome of interest will be the direction of hazard ratios seen using BRATECA and CRIS datasets. Therefore, validation analysis will be conducted in two stages: 1) analysis in BRATECA dataset, 2) analysis in CRIS dataset, and 3) comparison of hazard ratio to MIMIC-IV Clinical Dataset.

#### Analysis in BRATECA or CRIS

Analysis will be conducted using the BRATECA dataset with a Cox regression consistent with the test data set (Stage 2). We acknowledge the likely underpowered nature of the CRIS and BRATECA data sources. Thus, we are looking for similar treads towards to offer proposed evidence of validation.

We note that upon first inspection, BRATECA does not feature diagnoses, hence we will use time of first medication administration to time of first antipsychotic as proxy for admission time to event time.

If the direction of hazard ratios are consistent across datasets, then we will conclude that patients who are prescribed our target medication(s) may be protective.

## 6 Study Setting

MIMIC-IV is a publicly available database containing deidentified electronic health records from over 300,000 patients admitted to the Beth Israel Deaconess Medical Center (BIDMC), a Harvard Medical School teaching hospital in Boston, USA, between 2008-2019. The data will be accessed via Google BigQuery and downloaded to a King’s College London device.

BRATECA includes 73,040 admission records of 52,973 unique patients extracted from ten hospitals located in two Brazilian states, collected between 2020 and 2021. The data will be downloaded to a King’s College London device.

CRIS is a case register system that provides de-identified information from electronic clinical records relating to secondary and tertiary mental health care services from an NHS mental health trust that provides secondary mental health care to a population of over one million residents of South London. Referrals are accepted from general practitioners, specialist physicians, consultant psychiatrists, and community mental health teams. As is the system in the NHS in England and Wales, all referrals need to be approved by the local clinical commissioning groups before they can be seen at the service. A mandatory condition of access to CRIS data is that patient level data remains within the SLaM network at all times. In practice, this means that data extracted from CRIS for a project is not saved on a personal or non-SLaM laptop or an external hard drive or emailed out of the Trust. All analyses of patient level data must take place within SLaM.

## 7 Ethical and Regulatory Considerations

### Assessment and management of risk

Only people who have signed data use agreement on PhysioNet for MIMIC-IV and BRATECA; only people who are approved by the oversight committee for the CRIS platform have access to the data.

Data security issues: Data will be stored on King’s College London computer systems. Identifiable patient information issues: Analysis will take place using anonymised records.

### Research Ethics Committee (REC) and other Regulatory review

MIMIC-IV data was collected as part of routine clinical care. It has been deidentified and transformed. It is available to researchers who have completed training in human research and signed a data use agreement. It was approved for research by the institutional review boards of the Massachusetts Institute of Technology and BIDMC, who granted a waiver of informed consent and approved the sharing of the research resource. For the present retrospective study, two authors (ML and MM) signed the PhysioNet Credentialed Health Data Use Agreement 1.5.0 for MIMIC-IV, on May 8 2024 and July 15 2024 respectively.

BRATECA was deidentified according to the Health Insurance Portability and Accountability Act (HIPAA) standards using structured data cleansing and date shifting. The data was collected as part of a research project developed with several hospitals in Brazil. All data sharing was approved by each hospital. Ethical approval to use the hospitals’ datasets in this research was granted by the Brazilian National Research Ethics Committee under the number 46652521.9.0000.5530. For the present retrospective study, data access was granted on PhysioNet by the authors of the BRATECA dataset on June 7, 2024 for ML. MM applied for access on July 15 2024 but at time of writing of this protocol was not granted access yet.

South London and Maudsley NHS Trust have established CRIS in 2008 to allow searching and retrieval of comprehensive, yet de-identified clinical information for research purposes with a permission of secondary data analysis, approved by the Oxfordshire Research Ethics Committee C (reference 08/H0606/71+5). This present retrospective study was approved by the CRIS oversight committee on June 5, 2024, as <u>Project 24-029.</u> ML, MM and EG are approved for data access.

### Regulatory Review & Compliance

CRIS users are requested to provide feedback on the progress of their study. We will have to answer to the CRIS Administrator for this purpose for a mid-year review after project approval and until project completion. If our study has experienced a delay of greater than 3 months in starting, the Oversight Committee may request that the application is resubmitted.

### Amendments

Amendments will be tracked with appropriate version-controlled protocol changes and stored on King’s College London (KCL) computer systems.

### Peer review

The protocol has been reviewed by the Chief Investigator (ML) and Senior Methodologist (BC) prior to being sent for assessment by Senior Clinician (PB).

### Protocol compliance

Accidental protocol deviations can happen at any time. They will be adequately documented on the relevant forms and reported to the Chief Investigator and Sponsor immediately. Deviations from the protocol which are found to frequently recur are not acceptable, will require immediate action and could potentially be classified as a serious breach.

### Data protection and patient confidentiality

All investigators and study site staff will comply with the requirements of the Data Protection Act 1998 with regards to the collection, storage, processing and disclosure of personal information and will uphold the Act’s core principles.

For MIMIC and BRATECA, data will be transferred from the PhysioNet environment to KCL devices. For CRIS, relevant output and analysis will be received and conducted on a SLaM device.

These data will be the joint responsibility of the Chief Investigator and Senior Methodologist. Data will be stored on computer systems with password protected folders. Analysis will take place using anonymised records. Patient level and hospital level identifiers will be removed.

Data will be held until 01-12-2024 or until the per permitted research has been completed whichever is sooner. Data is stored in accordance with GDPR and The Data Protection Act 018 .

### Access to the final study datasets

The final datasets will only be accessed by the study team completing the analysis.

## 8 Dissemination Policy

### Dissemination policy

The work will be disseminated via peer review journal submission, conference presentation, social media, and press release with embargo date.

### Authorship eligibility guidelines and any intended use of professional writers

Authorship will be decided between the study team based on contribution to the final work. No professional writers will be used.

## Supporting information

Supplementary Material Signature Page

## Data Availability

MIMIC-IV and BRATECA are available after credentialing and training on physionet Anyone can apply to use CRIS provided they meet the governance requirements set out in the CRIS Security Model.

https://physionet.org

https://maudsleybrc.nihr.ac.uk/facilities/clinical-record-interactive-search-cris/information-for-researchers/

