## Supplementary Material Signature Page for "Protocol for: A Simple, Accessible, Literature-based Drug Repurposing Pipeline"

The undersigned confirm that the following protocol has been agreed and accepted and that the Chief Investigator agrees to conduct the study in compliance with the approved protocol and will adhere to the principles outlined in the Declaration of Helsinki, the Sponsor’s SOPs, and other regulatory requirement.

I agree to ensure that the confidential information contained in this document will not be used for any other purpose other than the evaluation or conduct of the investigation without the prior written consent of the Sponsor
I also confirm that I will make the findings of the study publicly available through publication or other dissemination tools without any unnecessary delay and that an honest accurate and transparent account of the study will be given; and that any discrepancies from the study as planned in this protocol will be explained.

**Chief Investigator:**

Signature:

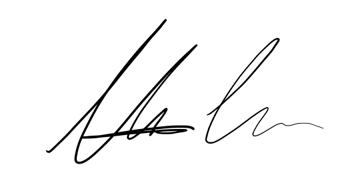

Date: 02/07/2024

Name: (please print):
Maximin Lange
